# The (mis-)alignment of genetic association studies to global health needs

**DOI:** 10.64898/2026.02.09.26345919

**Authors:** Rayan Alolayet, Amanda HW Chong, Robert W Aldridge, George Davey Smith, Gibran Hemani, Josephine G Walker

**Affiliations:** Executive Directorate of Pharmacovigilance, Drug Sector, Saudi Food and Drug Authority, Saudi Arabia; Population Health Sciences, Bristol Medical School, University of Bristol, United Kingdom; Medical Research Council Integrative Epidemiology Unit, University of Bristol, United Kingdom; Department of Health Metrics Sciences, University of Washington, Seattle, WA, USA

## Abstract

Health research priorities are generally not aligned with global disease burden. Although genome-wide association studies (GWAS) are correcting a historical bias by including samples from different demographic groups, this does not necessarily translate to improved understanding of the most important causes of disease globally. We demonstrate that while in countries with high socioeconomic development index (SDI) there is some alignment between the traits being analysed in GWAS and those that contribute most to disease burden, there is almost no such alignment in countries with low SDI. Improvement in alignment between GWAS and disease burden has been seen for countries with middle SDI over time, likely due to the contributions to disease burden changing in those regions rather than GWAS responding to the needs of those regions. Low GWAS alignment with disease burden may be partially explained by lower GWAS attention to childhood health. Improving aetiological understanding of high burden neglected conditions should be a priority for emerging biobanks in order to reduce global health inequality.

**Short abstract:** We identify some alignment between the traits being analysed in genome-wide association studies (GWAS) and disease burden in high socioeconomic development index (SDI) countries, while there is almost no such alignment in countries with low SDI, mostly due to neglecting childhood infection. Improvement in alignment between GWAS and disease burden has been seen for countries with middle SDI over time likely due to changing disease burden.

## Introduction

Health research priorities have been shown to be mis-aligned with disease burden, both at the national scale^1–3^, and globally^4^. Various efforts to coordinate global health research in alignment with health needs have been introduced in the 21^st^ century, but conflicting priorities for stakeholders including researchers, funders, publishers, and policy makers have limited efficient coordination of research funds^5^.

Genetic evidence is increasingly important for establishing effective health interventions. Over the past 20 years, thousands of genome-wide association studies (GWAS) have accumulated knowledge of biological and epidemiological mechanisms contributing to disease outcomes^6^. With a growing awareness that there is a major lack of ancestral diversity in GWAS^7^, it is now routine to involve samples from multiple ancestral backgrounds and to contrast or combine effect estimates across these demographic strata, though individuals who cluster with those of European ancestry still dominate multi-ancestry GWAS sample sizes^8^. There is an emerging trend that the marginal genetic effects typically estimated in GWAS are generally consistent across ancestral groups once accounting for technical artefacts and differential power^9^. As a result, evidence so far suggests that differential regional or global health inequalities cannot be substantially explained by differential genetic effects across populations.

However there is some emerging evidence that cross-population allele frequency differences of disease risk variants could explain a proportion of differential disease risk^8,10^, which motivates the continued expansion of diverse ancestral groups contributing to GWAS to improve transportability of genetic predictors. This observation invites the question of how else GWAS can contribute to improving health inequalities.

To this end we aim to evaluate if the coverage of disease burden studied through GWAS is representative of the disease burden experienced globally over 1990-2023. Disease burden is represented by the Global Burden of Disease (GBD) study which produces disability-adjusted life year (DALY) estimates combining morbidity and mortality by cause, broken down by time period, age group, sex, region, and socio-demographic index (SDI)^11^.

## Results

GWAS catalog traits were systematically mapped to 308 disease terms in the 2023 GBD database using a combination of manual and automated procedures by aligning with experimental factor ontology (EFO) terms (**Supplementary figure 1**). We excluded injury-related causes in GBD as they were unlikely to have a strong genetic component. Then, for every GBD disease term, we developed GWAS attention scores that use number of study participants, number of times that the EFO term was investigated, weighted number of independent GWAS hits, or weighted number of EFO by the impact factor of the journal as alternative measures to represent the degree to which a particular disease has been prioritised by the GWAS community. Note that all GWASs conducted over its 20 year history were aggregated into each score.

Firstly, using the Gini index calculated for 2023, we identify a substantial inequality in GWAS attention scores (Gini index = 0.94 **Figure 1A**). For interpretation, an index of 0 would indicate that GWAS attention was equal across traits, and as the index approaches 1 inequality increases meaning more attention is concentrated amongst fewer traits. This result is relatively consistent across different approaches to developing attention scores (ranging from 0.89 to 0.95, **Supplementary table 1**).

**Figure 1:**
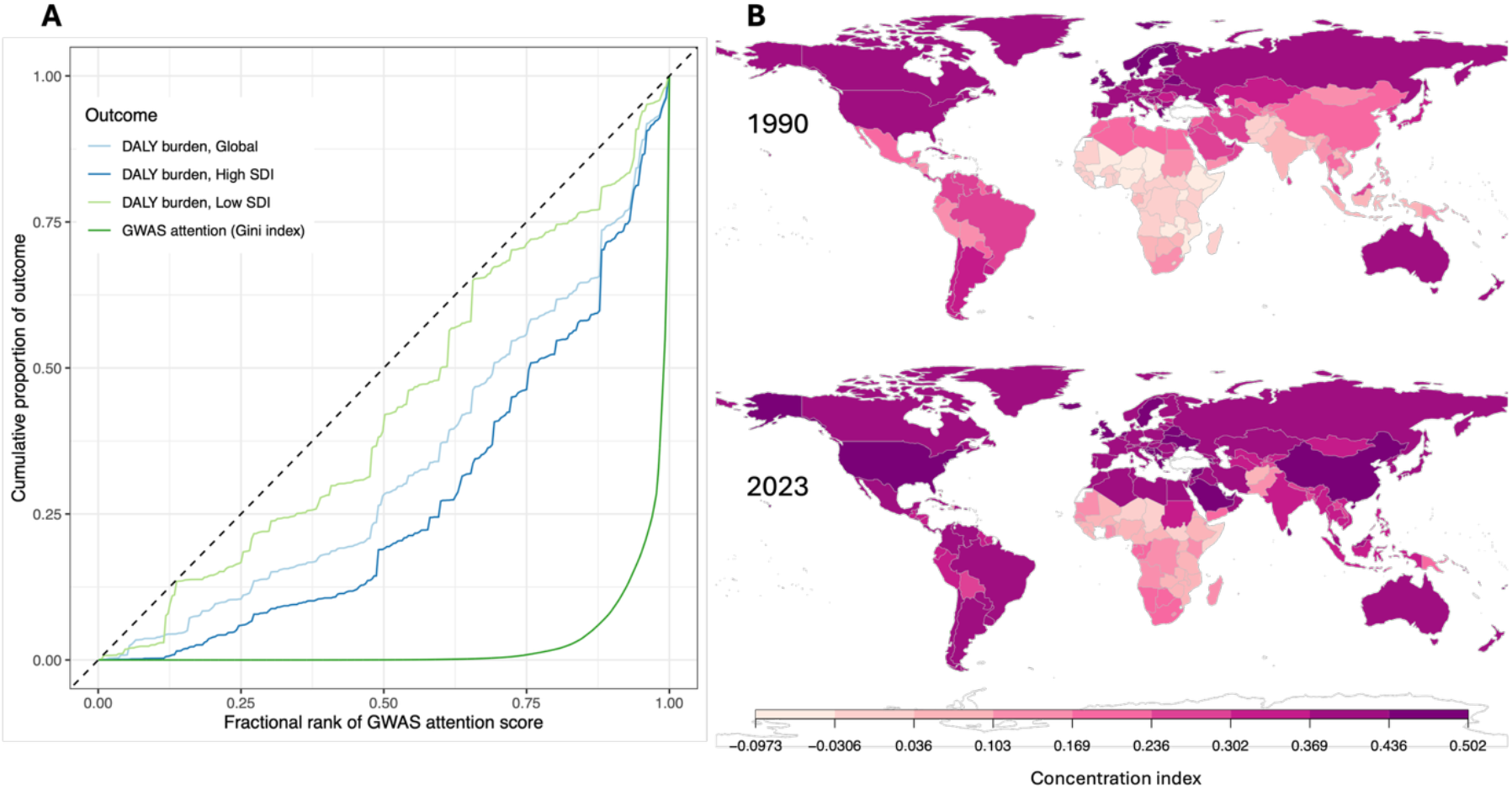
GWAS attention scores and DALYs. A) Lorenz curves illustrating the inequality in GWAS attention (dark green curve) illustrating that almost all attention is concentrated in few diseases. The alignment of these attention scores with corresponding DALYs are highest for DALY estimates from high socioeconomic index countries in 2023. B) The concentration index for each country is calculated based on the global GWAS attention scores alignment with that country’s DALY estimates. Visual comparison of alignment between 1990 and 2023 indicates that alignment is improving gradually but at different rates globally.

We next examined the extent to which GWAS attention scores align with the global DALY values attributed to each of those traits using a concentration index (CI). Under a scenario of perfect alignment of GWAS attention to global health needs as measured by DALYs, we would expect the CI to match the Gini index value of 0.92, whereas if there was complete randomness in terms of these two measures, we would expect a concentration index of 0; while a negative CI would indicate disproportionate concentration on the lowest burden diseases. We found a global alignment of 0.33 (standard error (se)= 0.06) in 2023. This indicates that there is a moderate correspondence of GWAS attention to global health needs. Stratifying DALY scores into country-level concentration indices we observe a more complex picture, with 147 of 204 countries analysed having a positive concentration index (where lower 95% confidence interval > 0). However, **Figure 1B** shows there is strong continental patterning, with particular misalignment of GWAS attention to health burden in Africa.

We hypothesised that socioeconomic development index (SDI) drives the difference in GWAS alignment to disease burden. This would arise for example if countries with higher SDI contribute more funding towards genetic studies, leading to data collection being centred within higher SDI regions, and hence biasing the representation of disease cases available for genetic studies. **Figure 2A** demonstrates that the concentration index drops substantially across SDI categories, with high SDI countries having relativelybetter alignment (0.45, se = 0.06), while low SDI countries show that the GWAS attention relates almost randomly to disease burden (0.14, se = 0.09) in 2019. The alignment of GWAS attention with disease burden appears to be improving over time, particularly in low-middle and middle SDI countries. We note that all SDI regions had a spike in concentration index estimates during the Covid-19 pandemic, owing to that being a large cause of disease burden globally at that time^12,13^ and also having substantial attention from GWAS. Comparison of concentration indexes across age groups consistently shows that inequality is largely driven by misalignment in children, especially in low and low-middle SDI countries (**Figure 2B**). Stratifying DALYs by sex led to relatively little difference in alignment of GWAS to disease burden (**Figure 2C**). Certain traits, such as diabetes mellitus, schizophrenia, and asthma, have received more attention than warranted by their global burden. On the other hand, neonatal disorders, lower respiratory infections, and diarrheal diseases have received particularly little attention relative to their burden.

**Figure 2:**
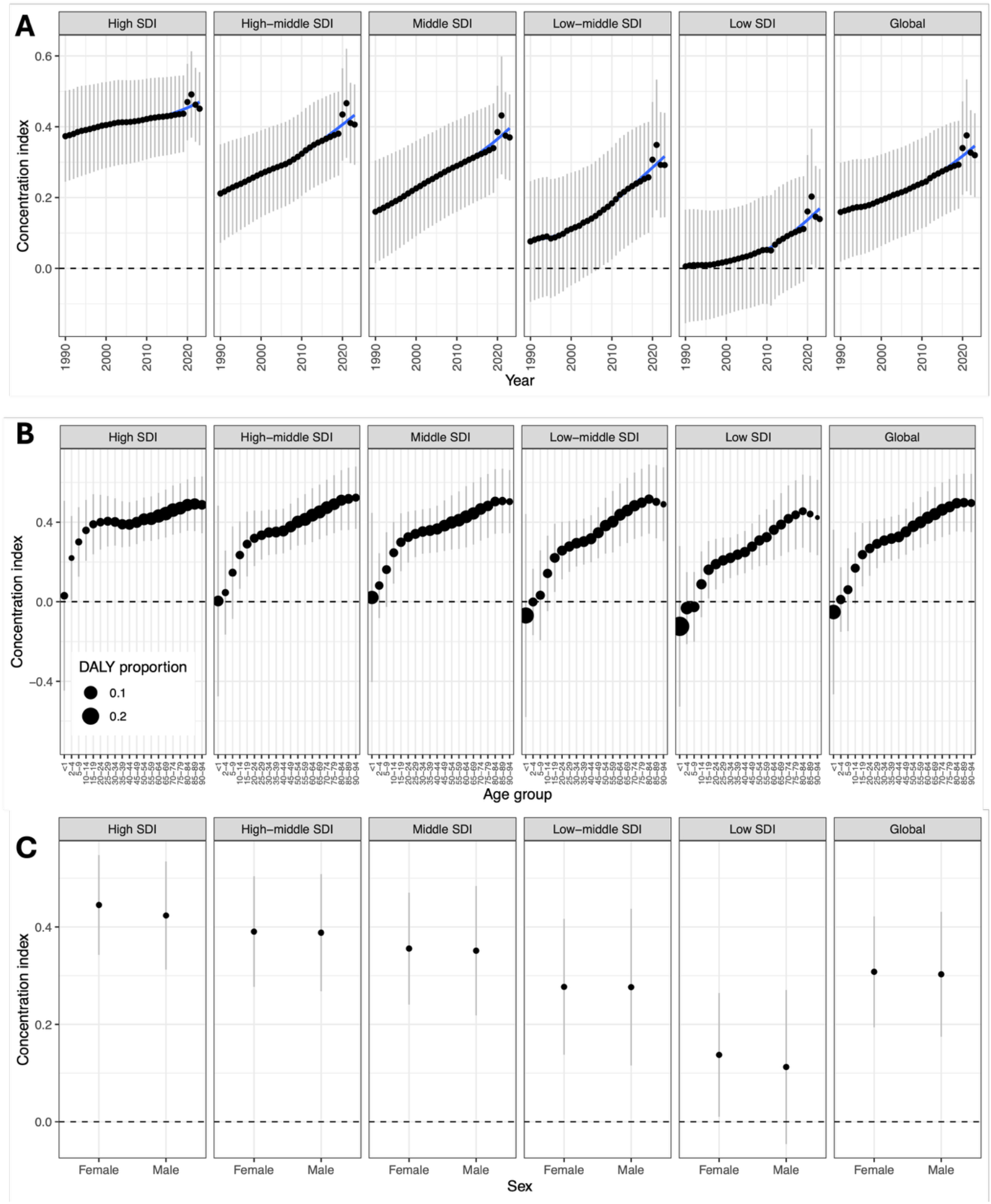
Demographic stratification of GWAS alignment to disease burden. Each point represents the concentration index with 95% confidence intervals. A) Concentration index (y-axis) stratified by SDI region (columns of plots) and estimated DALYs by year (x-axis). B) Differential disease burden by SDI category appears to be largely driven by lack of attention to childhood DALY contributions, because lower SDI countries have larger disease burden in children. C) Stratification of the concentration indexes where DALYs are stratified by sex.

## Discussion

In this study, we found that a small number of traits dominate GWAS efforts. This concentration suggests that research is disproportionately directed toward select conditions, while many traits receive minimal or no attention. Such skewness in the distribution may reflect structural factors, including the availability of large-scale biobank data for certain traits, historical research momentum where traits that were early targets of GWAS continue to attract attention, or perceived tractability of specific phenotypes. The repeated finding of high inequality across methods reinforces the robustness of this observation. Notably, this pattern parallels previous studies, such as Fitipaldi and Franks (2023)^14^, which showed that 40% of GWAS publications from 2005– 2022 focused on just 10 traits. This suggests an entrenched trend rather than a temporary artifact. While concentrated attention can drive rapid progress in select areas, it may also constrain the discovery of therapeutic targets, reinforce inequalities in research attention, and limit the development of a comprehensive understanding of human genetic architecture.

When we evaluated how well GWAS attention aligns with global health burden, measured via DALYs using the concentration index, we obtained a value of 0.33 (se = 0.06), indicating moderate alignment. This suggests that while some research focuses on high-burden traits, substantial discrepancies remain. Infectious diseases such as lower respiratory infections and diarrheal diseases, have received relatively limited GWAS attention despite their significant burden in many regions. It is important to note, however, that there is no argument against using GWAS to understand infectious disease^15,16^ as, during the COVID-19 pandemic, one of the largest GWAS ever was conducted relating to COVID-19 infection and outcomes^17^, demonstrating the ability of genetic studies to inform disease outcomes and guide public health responses. Indeed, (**Figure 2A)** shows that alignment between GWAS attention and disease burden increased in all regions during the COVID-19 pandemic.

Our analysis shows a clear decreasing trend in GWAS alignment from high to low SDI regions (**Figure 2A**). Although sample sizes may be small for many region-specific diseases, particularly those outside of European ancestry contexts, this should not preclude their inclusion. Innovative methods exist to help boost statistical power for such studies; various approaches^18,19^ can be used to leverage shared genetic architecture across related traits. The alignment between GWAS attention and the disease burden appears to be improving over time, especially in low-middle and middle SDI countries. However, this reflect a broader reduction in disease burden in these regions likely driven by improved access to healthcare and medical infrastructure^12^, rather than a strategic shift in GWAS priorities; the attention scores used here do not account for changes over time.

The misalignment indicated by concentration indexes across age groups shows that children in all SDI regions receive relatively less GWAS attention compared to their health burden (**Figure 2B**).Underrepresentation of pediatric diseases may present unique methodological and ethical challenges in genetic research.

There were a number of limitations in this study. Using the EFO terms enabled us to systematically integrate GWAS and GBD. However, the limitation of this approach is that if a trait in GWAS is mapped to an alternative EFO term, it could lead to categorising the EFO of that trait as unmatched, which may underestimate GWAS attention. Thus, we tried to minimise the implications of that by employing pattern and manual matching for unmatched terms. Another limitation is that there is variation in reporting the number of participants involved in GWAS among different studies. Hence, we utilised several approaches to measure the attention. When comparing different approaches, no considerable difference was observed.

GWAS has a role to play in reducing existing health inequalities, and being applied and developed in such a way as to not exacerbate existing health inequalities. As efforts continue to expand the global diversity of GWAS participants, examining diseases with the greatest regional importance will be a key factor in delivering the benefits of genetic studies to previously underserved communities.

## Methods

### Data sources

#### Global burden of disease study

The Global Burden of Disease Study was used to provide a comprehensive understanding of the health outcomes in the world^11^. It includes 375 diseases and injuries classified into a five-level hierarchy (0 represents all causes, and 3 and4 are the most detailed levels). Those diseases are divided into three broad categories, which include non-communicable disease (NCD) Communicable, Maternal, Neonatal, and Nutritional disease (CMNN), and injuries. We used DALYs to estimate the contribution of each disease to overall disease burden. Per-disease DALY estimates were obtained by country, within different SDI regions, by sex and age groups, and at multiple time points (from 1990 to 2023) using the IHME Global Health Data Exchange. From the GBD database we exported all most detailed causes (n=309, excluding causes at level 0, 1,2, and 3 which are more precisely captured at level 3 or 4) and thenexcluded all 30 remaining causes in the injury category (**Supplementary table 2**), to provide specific insights on genetic conditions that are more likely to be conducted by GWAS.

#### GWAS Catalog

The GWAS Catalog^20^ contains all association studies of the genetic variance linked to traits sourced from PubMed. We took the dataset of GWAS hits from the June 2024 release. It included 107,512 mapped traits to the EFO terms and other metadata including sample sizes from over 6,900 papers published between 2008 and 2024.

#### Experiment Factor Ontology (EFO) terms

The Experimental Factor Ontology (EFO) terms^21^ was used to map GBD diseases and integrate them to the EFO annotated GWAS traits. This hierarchical representation of terms standardized disease representation and facilitated data integration. It includes a comprehensive set of 260 ontologies with a total of 8,382,915 classes. The EFO terms were accessed via the Ontology Lookup Service on 15 June 2024.

#### Clarivate Journal Citation Reports (JCR)

To contribute to the estimation of GWAS study impact we obtained Clarivate Journal Citation Reports (JCR) based on the release from October 2023^22^. The GWAS Catalog includes 856 unique journals that published GWAS studies from 2008 to 2024. The JCR provides bibliometric analysis demonstrating the quality of each journal.

### Mapping GWAS and GBD terms

Integrating data from multiple sources specifically traits in GWAS, and health conditions in the GBD study is a complex process due to the two databases using different trait ontologies. We opted to map EFO terms to GBD terms, such that GWAS studies could then be clustered within GBD terms based on the GWAS study’s EFO term annotation. We initially manually searched for EFO terms aligning to the included GBD conditions in the NCD and CMNN categories. Subsequently, matching was performed between manually mapped EFO terms from the GBD and the EFO terms of traits in the annotated GWAS. This followed several steps to ensure consistent formatting, cleaning, and standardization of EFO terms. However, restricting the matching process to traits explicitly listed in the GWAS Catalog via EFO terms may underestimate GWAS attention due to the potential misalignment between traits and their EFO terms, as well as conditions that remained unmatched due to semantic differences. Therefore, pattern matching was conducted using the stringr^23^ package in R between GBD conditions and GWAS traits. For GBD conditions that remained unmatched due to semantic discrepancies, manual review of GWAS trait mappings were performed to improve alignment with GBD conditions. We utilized the OntologyIndex package in R to obtain descendants for those mapped EFO terms, if available. An overview of the process is provided in **Supplementary figure 1**.

### GWAS attention scores

To measure the attention of the GBD health conditions given by GWAS, we developed GWAS attention scores for each of the 274 GBD terms utilizing four approaches described in **Supplementary Table 1** in order to examine if our overall results were sensitive to the method of estimating GWAS attention. In the matching process, if there was more than one of EFO term matched with one GBD condition, the attention scores were added together as long as they arose from independent PubMed studies. This ensures that scores reflect the total attention received by their causes.

### GWAS inequality and alignment to disease burden

We measured the inequality of GWAS attention scores using the Gini index. We examined the extent to which GWAS attention scores align with the global DALY values attributed to each of those traits using the concentration index measure. The alignment of GWAS attention to global disease burden was also investigated across time (1990-2023) to assess whether research priorities have become more closely aligned with actual health needs over time. Additionally, we performed sex-stratified analyses and age group stratification to explore whether these alignments differ across demographic groups. We further stratified DALY scores into country-level concentration indices by SDI regions, as we hypothesized that differences in socioeconomic development drive variations in GWAS alignment to disease burden. To identify traits that were over- or under-attended by GWAS we regressed GWAS attention scores against DALY estimates and identified the disease outcomes that had largest positive and negative residuals.

## Data availability

All data is publicly available as described in the Methods, and harmonised and integrated versions of the data are available at https://github.com/Rolayet/GWAS-GBD

## Code availability

All code used to generate these results are available at https://github.com/Rolayet/GWAS-GBD

## Acknowledgements

GH, AHWC and GDS are funded by NIHR Biomedical Research Centre at the University Hospitals Bristol and the UK Medical Research Council Integrative Epidemiology Unit MC_UU_00032/1, MC_UU_00032/03. GH and GDS conduct research at the NIHR Biomedical Research Centre at the University Hospitals Bristol NHS Foundation Trust and the University of Bristol.

## Supplementary Figures

**Supplementary figure 1:**
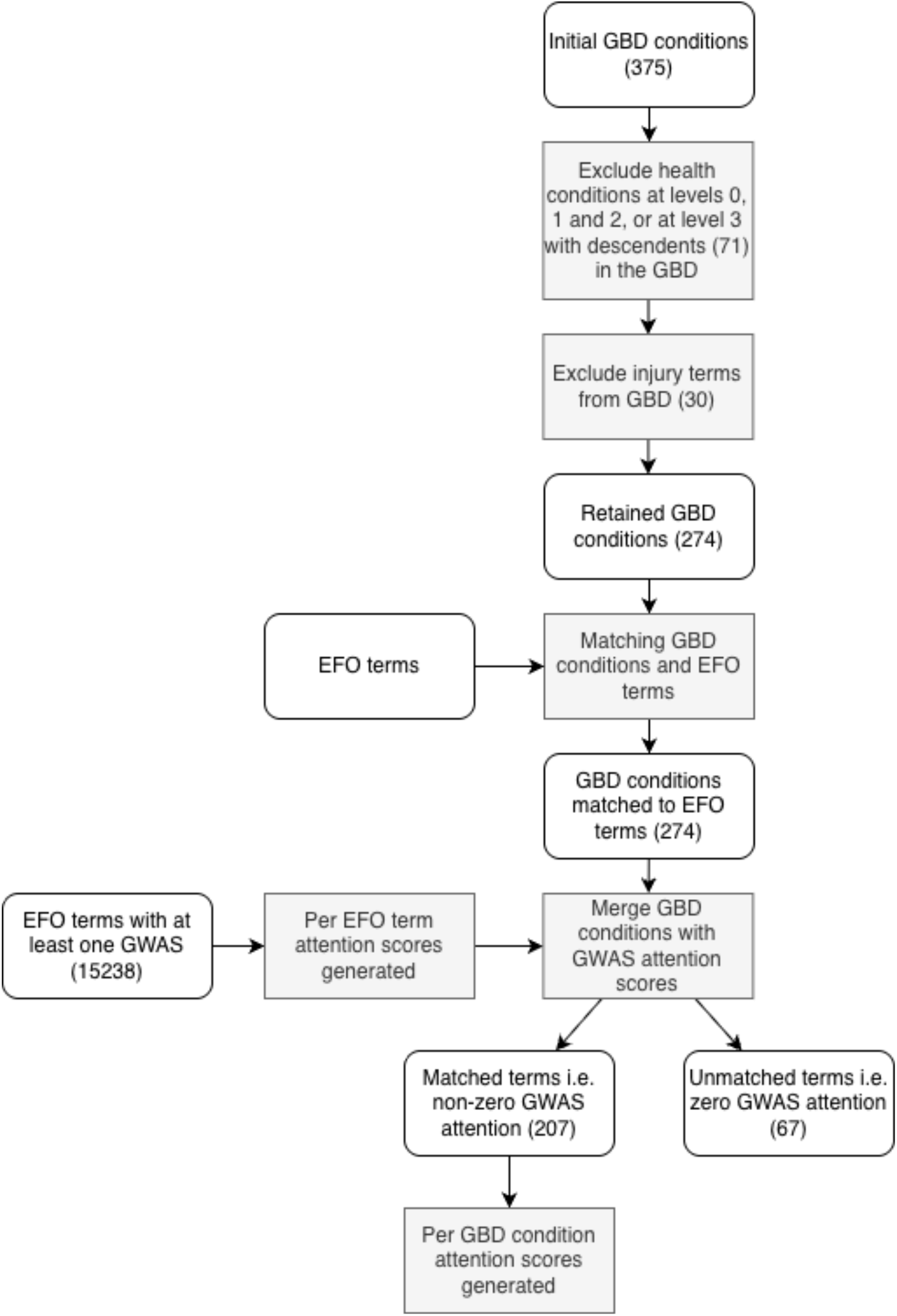
Flow chart showing the process of matching GBD conditions to GWAS traits.

## Supplementary Tables

**Supplementary Table 1:** Comparison of Gini index results among different GWAS attention scoring approaches.

| Approaches | Methods of Score Generation | Gini index |
| --- | --- | --- |
| **1. Number of participants** | In continuous traits, all participants were involved, while in binary traits, this was limited to the number of cases | 0.93 |
| **2. Number of studies** | Number of instances of the EFO terms | 0.89 |
| **3. Study impact factor** | As in (2) but studies were weighted by the impact factor of the journal | 0.95 |
| **4. GWAS hits** | As in (2) but studies were weighted by the number of independent GWAS hits reported | 0.93 |

**Supplementary Table 2:** Injury-related causes (category C) that were excluded from the analyses because they are unlikely to have a strong genetic aetiology. The list of all most detailed GBD causes was retrieved from the 2023 Global Burden of Disease (GBD) database.

|  |
| --- |
| Pedestrian road injuries |
| Cyclist road injuries |
| Motorcyclist road injuries |
| Motor vehicle road injuries |
| Other road injuries |
| Other transport injuries |
| Falls |
| Drowning |
| Fire, heat, and hot substances |
| Poisoning by carbon monoxide |
| Poisoning by other means |
| Unintentional firearm injuries |
| Other exposure to mechanical forces |
| Adverse effects of medical treatment |
| Venomous animal contact |
| Non-venomous animal contact |
| Pulmonary aspiration and foreign body in airway |
| Foreign body in eyes |
| Foreign body in other body part |
| Electrocution |
| Environmental heat and cold exposure |
| Exposure to forces of nature |
| Other unintentional injuries |
| Self-harm by firearm |
| Self-harm by other specified means |
| Physical violence by firearm |
| Physical violence by sharp object |
| Physical violence by other means |
| Conflict and terrorism |
| Police conflict and executions |

